# Insights on recurrent and sequential *Clostridioides difficile* infections from genomic surveillance in Minnesota, USA, 2019-2021

**DOI:** 10.1101/2025.05.13.25327503

**Authors:** Daniel Evans, Bree Friedman, Kelly Pung, Bonnie Weber, Matthew Plumb, Jacob Garfin, Christine Lees, Stacy Holzbauer, Ruth Lynfield, Xiong Wang

## Abstract

**Background:** The frequent temporal recurrence of *Clostridioides difficile* infection (CDI) may be the result of relapse with the same strain or reinfection with a different strain; the role of genetic evolution of *C. difficile* in relapse is poorly understood. We used whole-genome sequencing (WGS) to determine the frequency of same strain relapse among CDI recurrences and changes in the genetic diversity of individual strains responsible for relapses over time.

**Methods:** We analyzed data from active population- and laboratory-based surveillance of CDI in Minnesota, USA. We performed WGS on isolates collected from 306 patients with multiple CDI events during 2019-2021. We identified multi-locus sequence types (MLSTs), nucleotide variants, and putative mobile genetic elements (MGEs) from WGS data to study the genetic similarity and evolution of those *C. difficile* genomes.

**Results:** Among patients with multiple CDI events in the total surveillance period, 198 (64.7%) had multiple infections of the same MLST. Of these patients, 17.6% had multiple same-MLST CDI events >8 weeks apart. Among 232 temporally defined cases of recurrent CDI, 155 (66.8%) involved isolates of the same MLST. Among same-MLST isolates, there were no statistically significant correlations between accumulated mutations and elapsed time between CDI events. Analysis of sequential same-MLST *C. difficile* genomes showed evidence of gain or loss of putative mobile genetic elements (MGEs) in 45.6% of genome pairs.

**Conclusions:** Leveraging the largest CDI genomic dataset to date, our results confirm prior findings that recurrent CDI is a combination of reinfection and/or change in the ascendant strain in mixed infection, and relapse, while expanding knowledge on the evolution of pathogenic *C. difficile* strains in the human gastrointestinal tract.

## INTRODUCTION

*Clostridioides difficile* imposes an urgent and expanding burden on healthcare infrastructure. In the United States, incidence rates of *Clostridioides difficile* infection (CDI) exceed 100 per 100,000 persons [1], causing nearly 13,000 deaths and approximately $1 billion USD in attributable healthcare costs each year [2]. A key challenge in treating CDI lies in its opportunistic infection onset in patients with altered gut microbiota from antimicrobial therapy, who then must be treated with further antimicrobial therapy as their infection progresses [3]. Conceptually, recurrent infections may consist of relapses, caused by the persistence of a single strain of *C. difficile*, or reinfection, resulting from a new infection from a newly encountered strain [4]. In the United States, 15 to 40% of effectively treated incident CDIs are followed by recurrent infections, with many patients experiencing multiple recurrence events [4].

A commonly used surveillance definition for recurrent CDI accounts for an 8-week period of potential microbiome disruption following antimicrobial therapy [5]. This case definition, based primarily on clinical practice and patient monitoring following CDI events, was developed before the contemporary era of affordable whole-genome sequencing (WGS) for microbial pathogens [5]. Since its development, WGS has expanded insights into potential clonality, incidence of monoclonal versus polyclonal infections, and evolution of the *C. difficile* genome that collectively highlight the complexity of microbial pathogenesis that occurs in recurrent CDI [6]. Ribotyping, targeted sequencing, and WGS studies of *C. difficile* have yielded numerous insights into its evolution, transmission, colonization, and mechanisms of virulence [7–10]. These findings have shown that relapse events occur both within and beyond the temporal window of the recurrent CDI case definition [7–10].

Using WGS to study recurrent CDI may yield additional insights into the evolution of *C. difficile* during human infection [11]. Experiments on mutation rates have shown that *C. difficile* accumulates point mutations and other variations in nucleotide sequence more slowly than many other healthcare-associated bacterial pathogens [12]. Genomic investigations of *C. difficile* outbreaks therefore employ more stringent sequence identity cutoffs when assessing clonality of, or epidemiological associations among, infections [12–14]. Detailed genomic studies of multiple CDI events in the same patients can contextualize these findings, as those strains are subject to selective pressures imposed by the human immune system and antimicrobial therapies employed as tertiary prevention during outbreaks [15].

This study had two objectives. The first was to use a large genomic dataset of systematically collected pathogenic *C. difficile* isolates to examine sequential infections in the context of a commonly used, temporally based case definition of recurrent CDI. The second was to examine WGS data from those sequential isolates to strengthen the field’s understanding of *C. difficile* evolution, both by quantifying nucleotide mutation events over time and by gauging the incidence of gene gain and loss via horizontal transfer.

## METHODS

### Collection of patient clinical data

Minnesota participates in *C. difficile* surveillance through the Centers for Disease Control and Prevention’s (CDC’s) Emerging Infections Program (EIP) [1]. Since 2009, Minnesota has conducted active population- and laboratory-based surveillance in 4 counties (Stearns, Benton, Morrison, and Todd), with a fifth (Olmsted County) added in 2011 [1]. An incident CDI case was defined as the first positive *C. difficile* toxin or molecular assay on a stool specimen from a person at least 1 year of age during the study period [1, 5]. A recurrent CDI case was defined as an additional positive test between two and eight weeks (14 to 56 days) following an incident infection [5]. Any additional infection occurring more than 8 weeks after a prior infection in the same patient was classified as a new incident case for that patient. Specimen collection dates for recurrent CDI cases – as well as any CDI events occurring beyond the 8-week temporal limit – were documented throughout the three-year study period. Institutional review board approval was not necessary, since this study was conducted as a component of public health surveillance in the state of Minnesota, USA, subject to Minnesota Reporting Rules.

### Specimen collection and whole-genome sequencing

Specimens from *C. difficile*-positive cases were transported to the Minnesota Public Health Laboratory, cultured on Taurocholate-Cefoxitin-Cycloserine-Fructose agar (TCCFA), and incubated anaerobically at 35°C for 48-72 hours. From each culture, a single suspected colony of *C. difficile* was sub-cultured to a blood agar plate. Cultures grown from that incubation step were then evaluated for bacterial species using the Bruker MALDI-TOF (Matrix-Assisted Laser Desorption ionization-time of flight) instrument using methods described previously [16]. DNA was extracted from each isolate using the Qiagen QIAamp BiOstic Bacteremia DNA Kit. DNA extracts were quantified on the SpectraMax M2 with the Invitrogen Qubit 1X dsDNA High Sensitivity Assay Kit and prepared for sequencing using the Illumina DNA Prep Kit (Illumina, Inc.). Sequencing was then performed using the Illumina MiSeq with the 500 cycle V2 kit or the Illumina NextSeq platform with the 300 cycle P1 cartridge. Long-read whole-genome sequencing was performed on selected isolates from the same DNA extracts used for short-read sequencing, using the GridION platform (Oxford Nanopore Technologies) and R9 sequencing chemistry.

### Analyses and comparisons of whole-genome sequencing data

Quality assessment of sequencing reads from 743 isolates collected from 306 patients was performed using FastQC v0.11.8 [17]. Species identification and contamination detection were performed using Kraken2 v2.0.8 [18]. Genomes were assembled from reads using Shovill v1.0.4 [19], and quality assessment of assemblies was performed using QUAST v5.0.2 [20]. Multi-locus sequence typing (MLST) was performed using MLST v2.17.6 [21, 22]. Genomes that showed no evidence of mixed strain contamination or erroneous base-calling during genome assembly were then annotated using Prokka v1.14.6 [23]. Core-genome alignments and pangenome data were generated using Roary v3.13.0 [24]. Maximum-likelihood phylogenetic trees were constructed from core-genome alignments using IQTree v2.3.3 [25]. Pairwise variant calling was performed using GBK files generated by Prokka and FASTQ sequencing reads using Snippy v4.6.0 [26]. Hybrid genome assembly from short-read and long-read data was performed using Unicycler v0.5.0 [27] and Flye v2.9.1-b1780 [28].

### Detection of putative mobile genetic elements

Identification of putative fragments of mobile genetic elements (MGEs) within non-contiguous, short read-sequenced genomes was performed using ABRicate v.1.0.1 [29]. Briefly, ABRicate was used to construct and search against custom reference databases of plasmid replicons from PlasmidFinder, as well as 2-kilobase fragments of MGE sequences collected from the MobileElementFinder v1.0.2 and ICEberg 2.0 databases [30, 31]. Comparisons of the presence and absence of annotated genes were performed using the “query_pan_genome” function of Roary v3.13.0 [24], to search for putative MGEs gained or lost by sequential pairs of same-MLST, same-patient isolates. Briefly, this function was used to identify contigs with at least one annotated open reading frame with a function related to mobilizing DNA (transposases, replicative helicases, plasmid partitioning, etc.) and at least two other open reading frames with any annotation, all of which were not found in the other genome in the pair. Confirmation of the presence or absence of putative MGEs was performed on hybrid genome assemblies using the mob-suite v3.0.3_lite package integrated into Flye v2.9.1-b1780 [32].

### Statistical analysis of variant data

Simple and multiple linear regressions of nucleotide variants accumulated over time within same-MLST infections in the same patients were performed using R statistical software v4.3.1 [33]. Sensitivity and robustness analyses were performed using sensemakr version 0.1.4 [34]. For analyses of nucleotide variants accumulated over time, statistical outliers in both elapsed time and variants identified were removed using the 1.5 interquartile range rule.

## RESULTS

### Broad genetic diversity among *C. difficile* strains that caused multiple infections

Within the three-year sampling period from January 2019 to December 2021, 306 patients had two or more CDI events documented (n = 743 total infections) (Figure 1). WGS of isolates from these patients confirmed that 198 (64.7%) patients had stool specimens from which we isolated at least two *C. difficile* strains of the same MLST. Of those patients, 165 (83.3%) had specimens from which only strains of the same MLSTs were isolated, and 33 (16.7%) had at least three infections of multiple MLSTs.

**Figure 1:**
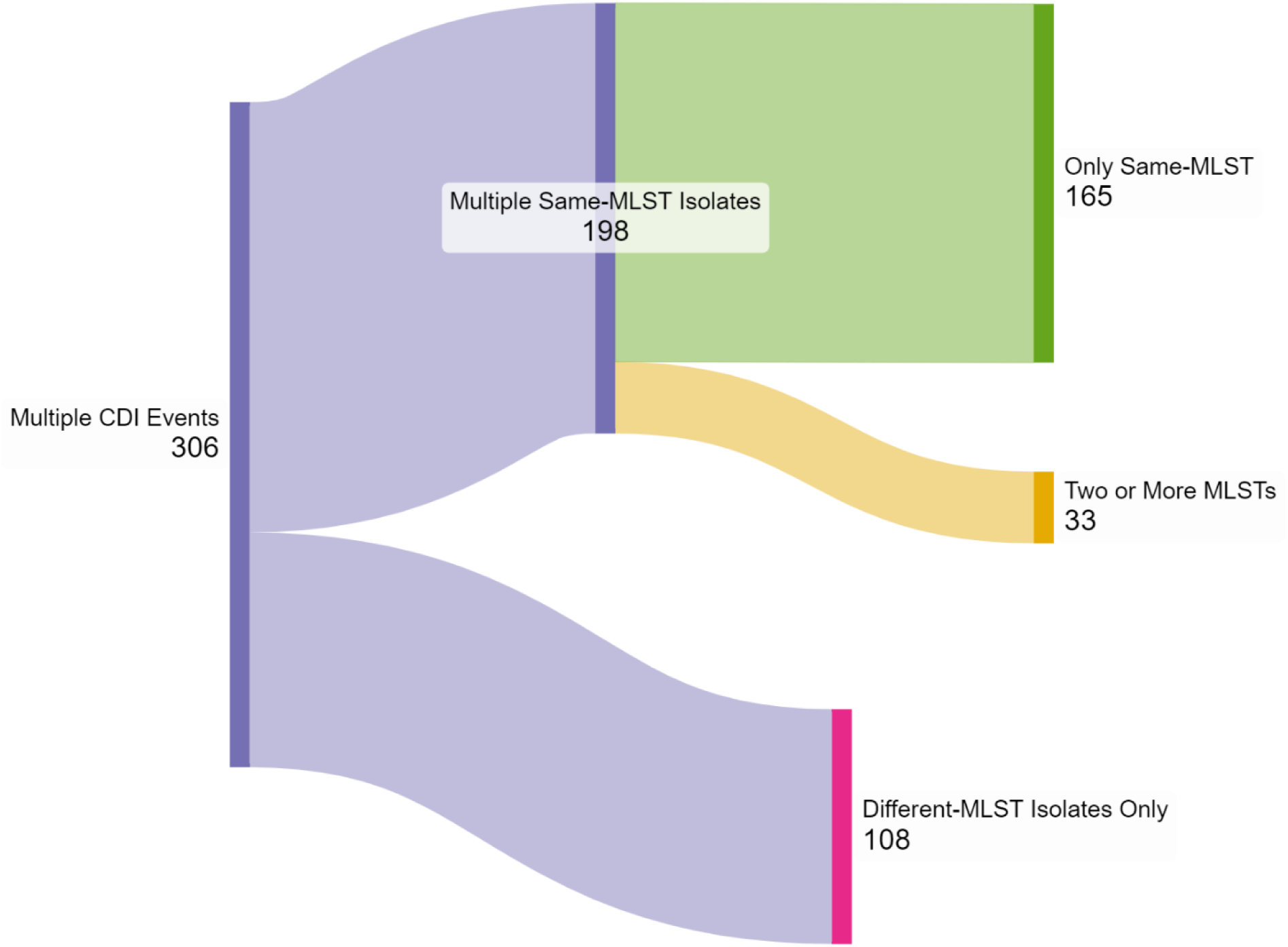
Sankey diagram showing the distribution of patients infected with multiple *C. difficile* infections during the three-year surveillance period, by isolation of same-MLST versus different-MLST strains. Numbers represent patients within the dataset. Figure generated from surveillance data using SankeyMATIC software (https://sankeymatic.com).

Within our dataset of multiple infections in the same patients, 501 of 743 (67.4%) isolates were from patients who met the temporally defined criteria for recurrent CDI, described earlier [1, 5]. These isolates comprised 232 events of one or more recurrent infections among 221 patients. MLST profiling of WGS data from these infections showed that of the 232 recurrent events, 155 (66.8%) involved isolates only of the same MLST, and 77 (33.2%) involved isolates of multiple MLSTs.

Focusing on high-quality *C. difficile* genomes from same-patient, same-MLST infections yielded 433 assemblies from 187 patients that could be further evaluated for specific genetic signatures of molecular evolution over sequential CDI events (Figure 2). These included genomes from 155 patients (82.9%) infected only by same-MLST isolates and from 32 patients (17.1%) infected by isolates of two or more MLSTs. These genomes were distributed across 39 MLSTs, the most abundant of which were STs 42 (n = 52, 12.0%), 2 (n = 44, 10.2%), 11 (n = 34, 7.9%), 8 (n = 29, 6.7%), 1 (n = 20, 4.6%), and 43 (n = 20, 4.6%) (Table 1).

**Figure 2:**
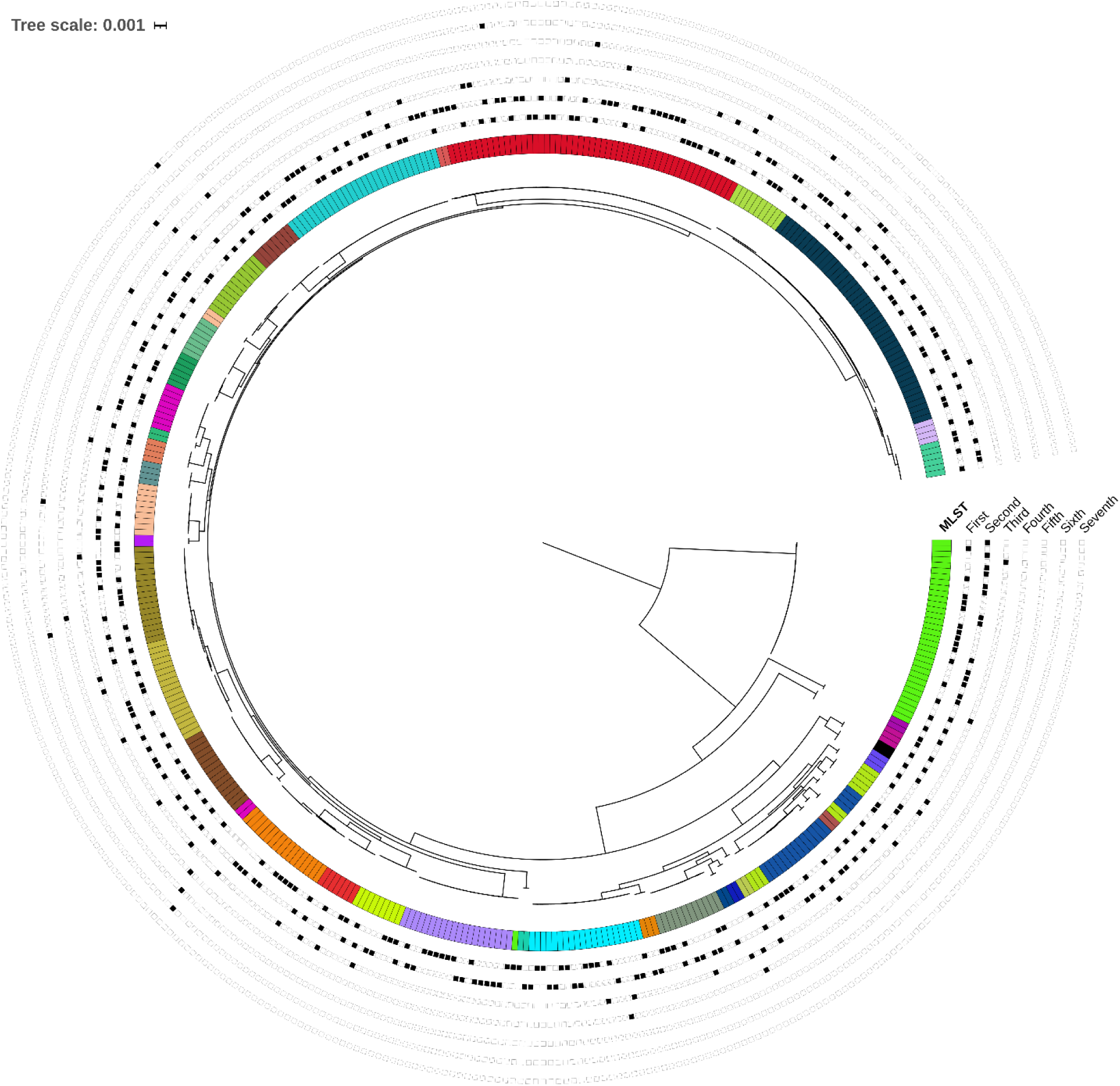
Midpoint-rooted maximum likelihood phylogenetic tree of 433 whole-genome sequences of *C. difficile* isolates collected from cases with multiple infections of the same multi-locus sequence types (MLSTs) during the two-year surveillance period. The tree was constructed using IQTree v2.3.3 from a core genome alignment of 2,571 genes, which was generated using Roary v3.13.0 [23, 24]. The color strip denotes MLSTs of each genome. Genomes are also numbered in the chronological order of cases’ same-MLST infections, with black squares annotating their place in the order of sequential infections.

**Table 1:** Summary of the multi-locus sequence types (MLSTs) of high-quality *C. difficile* genomes from patients with two or more same-MLST infections.

| *Multi-Locus*<br>*Sequence Type*<br>*(MLST)* | *Total Isolates* |
| --- | --- |
| 42 | 52 |
| 2 | 44 |
| 11 | 34 |
| 8 | 29 |
| 1 | 20 |
| 43 | 20 |
| 58 | 18 |
| 67 | 18 |
| 34 | 17 |
| 53 | 17 |
| 55 | 15 |
| 54 | 12 |
| 95 | 12 |
| 3 | 11 |
| 41 | 10 |
| 110 | 10 |
| 4 | 9 |
| 6 | 8 |
| 10 | 8 |
| 21 | 7 |
| 103 | 7 |
| 14 | 6 |
| 35 | 6 |
| 9 | 4 |
| 46 | 4 |
| 49 | 4 |
| 221 | 3 |
| 224 | 3 |
| 228 | 3 |
| 12 | 2 |
| 28 | 2 |
| 32 | 2 |
| 37 | 2 |
| 44 | 2 |
| 47 | 2 |
| 92 | 2 |
| 97 | 2 |
| 190 | 2 |
| 231 | 2 |
| Unclassified | 2 |
| **TOTAL** | **433** |

### Subsequent *C. difficile* infections caused by the same MLST occur more than eight weeks after initial infection

Of the 187 patients whose *C. difficile* isolate genomes we further evaluated for molecular evolution, 133 (71.1%) had subsequent same-MLST infections only within 2 to 8 weeks after incident infection, 36 (19.2%) only greater than 8 weeks, and 18 (9.6%) with at least one infection within 2-8 weeks and at least one infection beyond 8 weeks. Among these patients, third, fourth, and additional infections of the same MLSTs were much more likely to occur after a greater elapsed time following a prior infection, compared to elapsed time between first and second same-MLST infections (Wilcoxon rank-sum test: p < 0.001) (Table 2). Patients with two or more same-MLST events greater than 8 weeks apart comprised 54 (17.6%) of all 306 patients with multiple documented CDI events.

**Table 2:** Summary and statistical comparison of elapsed times in between sequential CDI events caused by isolates of the same MLSTs. Calculations were performed on 237 pairs of sequential isolates from the same patients, following removal of statistical outliers. Isolate pairs were divided into those including the first and second same-MLST infections only, and those including any additional infections of the same MLSTs at any point in the surveillance period. Comparisons were performed using the Wilcoxon rank-sum test for non-parametric data, using a significance threshold of α = 0.05.

| *Pairs of Sequential Infections of Same MLST* | *Number of Isolate Pairs* | *Mean Days Between Infections (Standard Deviation)* | *Median Days Between Infections (IQR)* | *P-Value* |
| --- | --- | --- | --- | --- |
| *First (Incident) and Second Infections* | n = 191 | 45.0 days<br>(32.4 days) | 35 days<br>(24 – 53 days) | P < 3 x 10<sup>-9</sup> |
| *Second and Third, Third and Fourth, etc.* | n = 46 | 79.4 days<br>(48.4 days) | 66 days<br>(48 – 103.5 days) |  |

### Mutation rates calculated from same-MLST *C. difficile* infections often exceed established nucleotide identity thresholds

To further investigate the evolution of *C. difficile* isolate genomes from CDI cases, we identified nucleotide variants – including SNPs and insertions and deletions (indels) – that accumulated in alignments of genomic DNA of sequentially collected same-MLST genomes from individual patients (Figure 3). Isolates accumulated a median of 1 total nucleotide variant (interquartile range 0-3 variants) and a median of 1 SNP (interquartile range 0-2.5 SNPs) in between symptomatic infections. Curiously, 24.9% of these pairwise comparisons differed by more than 2 SNPs, a value frequently used in genomic investigations of *C. difficile* transmission as a threshold to screen for outbreaks [14]. The median mutation rate – calculated by dividing total nucleotide variants by elapsed time between specimen collection – was 9.6 variants per year (interquartile range 0-30.6 variants/year). Neither accumulated variants nor accumulated SNPs in sequential pairs of same-MLST isolates were statistically significantly correlated with elapsed time between infections (Table 3). Sensitivity analyses established no significant robustness with respect to meeting case definition criteria for recurrence, nor to the number of previous same-MLST infections (Table 3).

**Figure 3:**
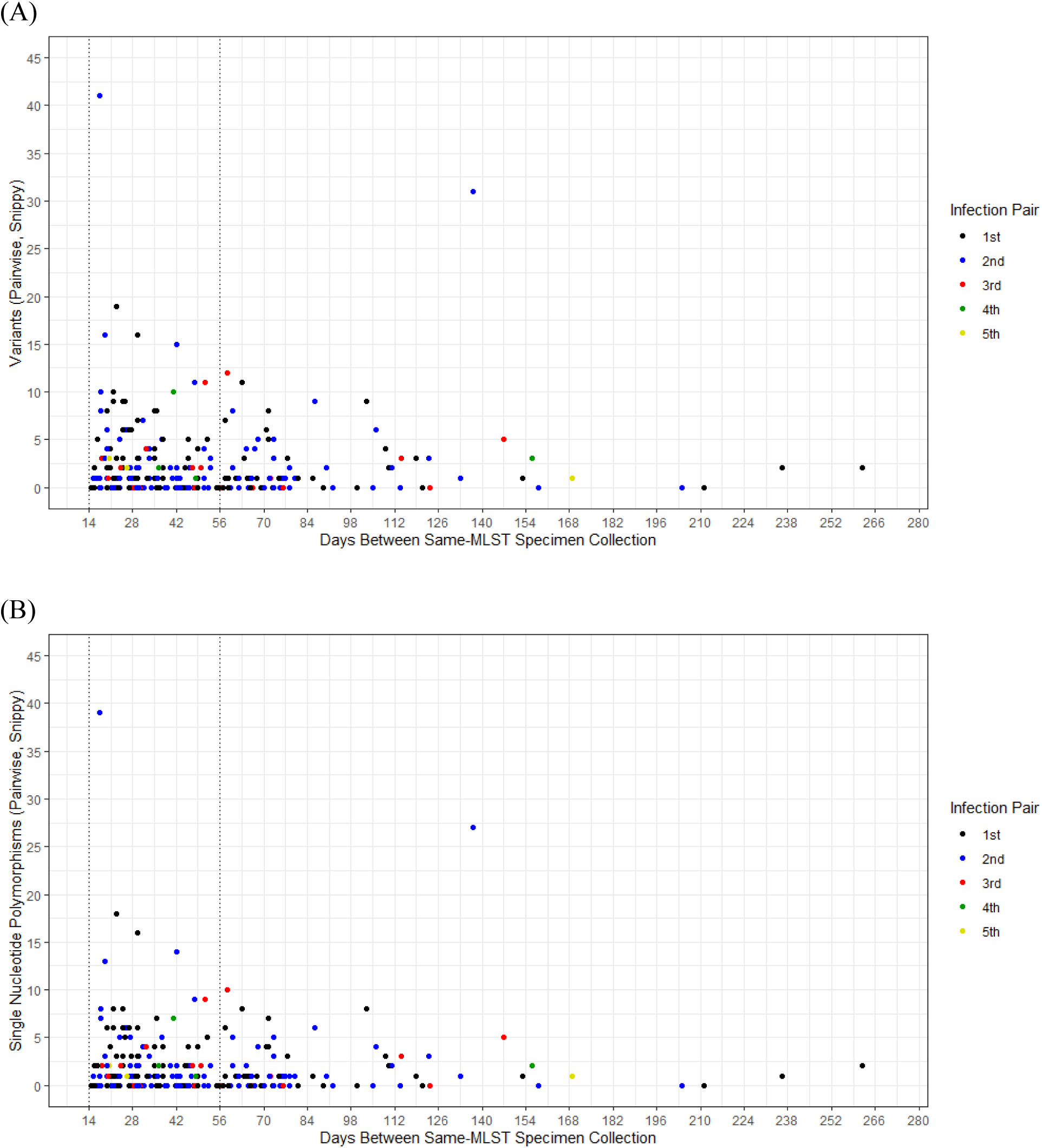
Scatter plots of mutations accumulated between same-MLST *C. difficile* infections in the same patients over time. Calculations were performed by pairwise alignments of sequencing reads of an isolate to the assembled whole-genome sequence of the previous isolate of the same MLST from the same patient. Panels show counts of (A) all nucleotide variants and (B) single-nucleotide polymorphisms (SNPs). Data points are colored by the sequential same-MLST isolates compared for each result; “1st” denotes the first and second isolate from the same patient, “2nd” denotes the second and third, etc. Vertical dotted lines show the temporal range of 14 to 56 days following prior infection, the key criterion to classify CDI as “recurrent”.

**Table 3:**
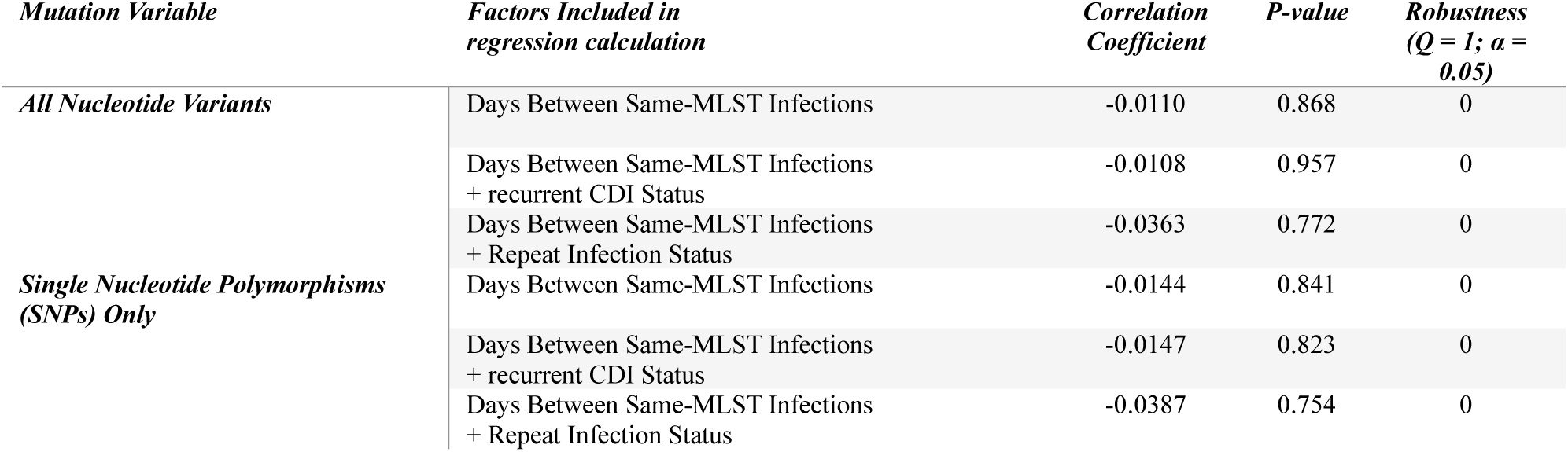
Summary of simple and multiple linear regressions of accumulated nucleotide variants over time among sequential pairs of *C. difficile* isolates of the same MLSTs collected from the same patients. Calculations were performed from variant calling of 237 pairs of isolates performed using Snippy v4.6.0 [25], following removal of statistical outliers for accumulated variants and time between infections. Non-synonymous nucleotide variants were counted if they occurred in annotated open reading frames and were predicted to affect the translated amino acid sequences of putative proteins. Days between infections were calculated from dates of specimen collection between isolates from infections separated by at least 14 days of elapsed time. Recurrent CDI (rCDI) status was included as a binary factor, denoting whether the second infection in each pair occurred less than 8 weeks after the first infection. Repeat infection status was included as a binary factor, denoting whether the isolate pairs were the first and second of the same MLSTs collected from each patient, regardless of elapsed time between infections. Correlation coefficients and p-values were calculated using R statistical software v4.3.1 [30], and robustness calculations were performed in R v4.3.1 using the sensemakr package [31].

### *C. difficile* strains gain and lose genetic cargo associated with putative mobile genetic elements in the human gastrointestinal tract

To expand our investigations into the evolution of *C. difficile*, we reviewed same-MLST genomes from individual patients for evidence of the gain and loss of putative mobile genetic elements (MGEs). We performed this by two approaches; in the first, we reviewed sequentially collected pairs of *C. difficile* genomes for the presence of different DNA sequences carried by known mobile genetic elements [29, 30]. In the second, we used Roary pangenomics software to compare sequentially collected genome pairs for differences in their carriage of annotated DNA mobilization genes adjacent to other open reading frames on the same contigs [23]. We supplemented these approaches with long-read sequencing and hybrid genome assembly [26, 27].

Our approaches showed that same-patient, same-MLST genomes collected frequently differed in genetic cargo associated with putative MGEs (Figure 4a). Among 248 pairs of same-MLST sequential isolates, 113 (45.6%) showed differences in the presence of putative MGE sequences by either approach. These included 76 pairs (30.6%) with evidence of gene gain and 69 pairs (27.8%) with evidence of gene loss. There were 32 pairs (12.9%) that showed evidence of both gain and loss of different putative MGE sequences. Based on substantial differences in their genetic cargo, five pairs of sequential same-MLST isolates were chosen for long-read sequencing and hybrid genome assembly. Two pairs showed the presence of additional contiguous mobile genetic element sequences in the chronologically earlier isolates, one showed the presence of an additional plasmid in the later isolate, and two showed the presence of additional plasmid sequences in the earlier isolates (Figure 4b).

**Figure 4:**
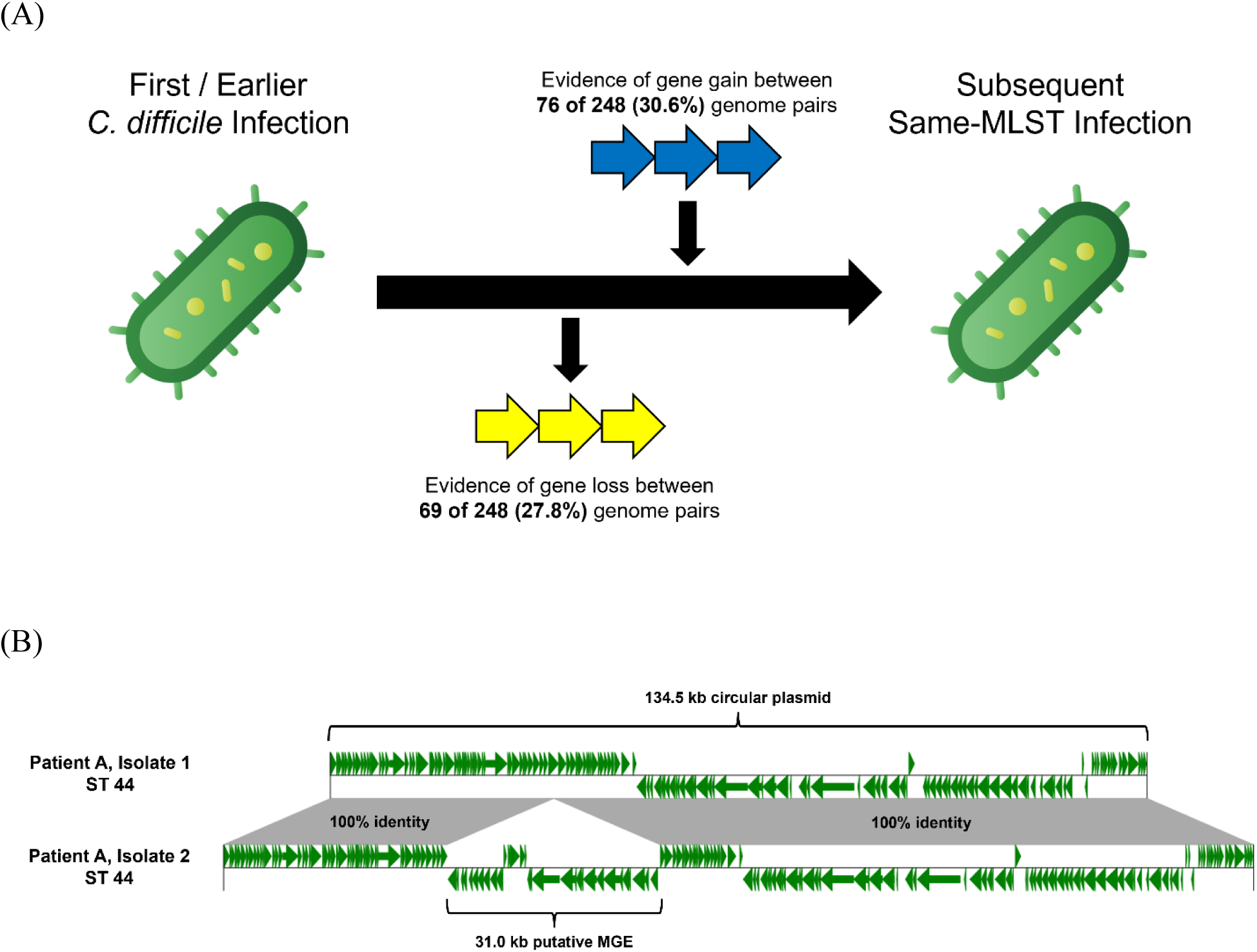
Gain and loss of genetic cargo associated with mobile genetic elements (MGEs) in between sequential same-MLST CDI and recurrent CDI events. (A) Diagram of putative gene flow in between same-patient, same-MLST infections, as detected by nucleotide alignment-based or pangenomic comparison-based approaches. Image was generated using graphics freely available from Flaticon (https://flaticon.com). (B) Alignment of a partial fragment sequence from two *C. difficile* isolates collected from the same patient 52 days apart, showing the presence of a 31.0 kb putative MGE in the chronologically later isolate. Both isolates were classified as MLST 44 and were otherwise genetically identical across the entirety of their shared genome sequences. Annotations genes in the contiguous segment unique to the second isolate included a tyrosine recombinase, excisionase, transposase, DNA topoisomerase, and bacitracin resistance ABC transporter machinery. The pairwise alignment figure was generated from hybrid genome assemblies with GBKViz (https://gbkviz.streamlit.app/).

## DISCUSSION

In this study, we applied whole-genome sequencing (WGS) to investigate C. *difficile* isolates that caused sequential infections in a large cohort of patients. We identified numerous events of strains of the same MLSTs causing multiple infections beyond the conventional temporal limits for defining recurrent CDI, as well as evidence that many recurrent CDI events involve isolates of different MLSTs than prior incident infections. We also found no evidence of a linear association between accumulated nucleotide variants and elapsed time between same-patient, same-MLST infections, but we observed the gain or loss of genetic cargo associated with putative mobile genetic elements (MGEs) in between sequential collections of *C. difficile* isolates.

Our findings highlight the complex microbial genetic diversity of strains involved in multiple events of CDI in the same patients and reinforce those of prior studies that used smaller genomic datasets [7–10]. By characterizing genomes through MLST and by identifying variants from pairwise genome alignments, we found many examples of recurrent infections within 8 weeks of prior infections that involved isolates of different MLSTs, and many others beyond 8 weeks that involved closely related strains. While interpreting these findings, it is critical to note they cannot be used as a direct quantitative measure to evaluate the sensitivity and specificity of temporally limited case definitions of recurrent CDI, with respect to the clonality of recurrent infections. Prior studies have repeatedly confirmed that CDI can be caused by multiple coinfecting strains, which can only be reliably assessed by laboratory or sequencing methods designed to assess the full genetic diversity of *C. difficile* strains collected from specimens [35–37]. However, our sequencing protocol involved the collection of single colony isolates from individual infections, thereby inherently limiting the assessment of genetic diversity within each confirmed CDI event. Prior findings that polyclonal infections likely constitute only a minority, not the majority, of CDI events reinforce our broader interpretations about the genomic complexity that temporally limited recurrent CDI case definitions fail to capture [35–37]. Until other studies employ methods that fully account for this complexity, our reported proportions of same-MLST versus different-MLST infections in the same patients should not be considered a definitive summary of the genetic diversity of recurrent and relapse CDI.

Additional questions arise from our results based on our methods for defining genetic similarity among *C. difficile* genomes. Investigating the frequency and phenotypic effects of mutations of CDI-causing strains as they cause multiple infections over time may yield useful insights into pathogenesis and intervention [7–10]. Prior studies have quantified genetic differences between genomes of isolates involved in recurrent or relapse infections by using MLST only or core-genome multi-locus sequence typing (cgMLST) [10]. These differences in bioinformatic methods complicate broader interpretations and increase the risk of inconsistency of findings from studying *C. difficile* infections at sub-species resolutions. Without uniform consensus on how to incorporate genomics methods into these analyses, epidemiological studies of CDI will continue to be limited by these inconsistencies. Until such consensus is reached, it is important for genomic studies to use caution in their terminology and use method-specific language as appropriate.

By including a preliminary investigation of mobile genetic elements (MGEs) in our study of recurrent CDI, we expand ongoing work to investigate the role of horizontal gene transfer (HGT) in the evolution of high-priority bacterial pathogens. Recent literature has demonstrated that these pathogens frequently exchange plasmids, transposons, and other MGEs [38–40]. Even though we employed both a DNA reference-based and a pangenomics-based approach to identify putative MGEs that were gained or lost among recurrent CDI-causing strains over time, we believe that our results underestimate the flow of genetic material among *C. difficile* during prolonged colonization and infection. Reference-based approaches that depend on extensive databases of MGE sequences cannot account for the presence or absence of those that have not been discovered or characterized [41, 42]. Similarly, our manual screening of annotated open reading frames in selectively present or absent sequences highlighted through our pangenomics approach likely filtered out many MGE sequences whose gene functions remain unknown [42, 43]. Longitudinal studies of sequential same-patient infections may serve as an ideal approach to further investigate how HGT influences evolution, pathogenesis, and virulence of high-priority bacterial pathogens [38, 40–43]. Such findings could better characterize MGEs’ effects on microbial evolution under selective pressures and on clinical outcomes of infected patients.

Despite its large quantity of genomic data and temporal scope of CDI surveillance, our study has several limitations. First, our use of WGS of single colonies isolated from cases’ stool specimens prevented investigation of polyclonal CDI events, potentially skewing our results on same-MLST versus different-MLST infections. Although we used long-read sequencing and hybrid genome assembly to investigate selected *C. difficile* isolate genomes, our use of short-read sequencing for most of our WGS data generation – assemblies from which frequently fragment MGE sequences into separate contigs – complicated our investigations into HGT and MGEs [44]. The geographical limitation of our genomic surveillance also potentially limited the generalizability of our results to other locations. Lastly, our methods for tracking HGT in between sequential CDI events were not designed to identify putative MGEs that lacked thorough documentation in reference databases or characterization by current genome annotation tools.

Our study highlights the value of incorporating WGS analyses in active surveillance of *C. difficile* to assess microbial diversity and evolution of this pathogen in scopes beyond outbreak investigations. Future studies should investigate microbial genomic and epidemiological factors that may affect the risk of sequential infections by the same *C. difficile* strains, apply metagenomic sequencing to characterize potential polyclonal infections, investigate non-synonymous mutations that emerge during sequential infections, examine the potential contributions of MGEs to recurrent CDI onset and severity, and compare WGS data from isolates that caused multiple infections to data from isolates that caused only single infections.

## Data Availability

All bacterial whole-genome sequencing data associated with this study have been uploaded to the NCBI website under BioProject number PRJNA629351.

https://www.ncbi.nlm.nih.gov/bioproject/629351

## ACKNOWLEDGEMENTS

We thank our collaborators at the Centers for Disease Control and Prevention (CDC) for their guidance throughout this project and their constructive feedback on this manuscript. We thank Clifford McDonald, Amy Gargis, Alice Guh, Alison Laufer Halpin, and Susannah McKay. The findings and conclusions in this report are those of the authors and do not necessarily represent the official position of the U.S. Centers for Disease Control and Prevention. We also thank members of the Clinical Microbiology unit of the Infectious Disease Laboratory at the Minnesota Department of Health for their support in processing, culturing, and analyzing *C. difficile* isolates prior to WGS. All bacterial whole-genome sequencing data associated with this study have been uploaded to NCBI under BioProject number PJRNA629351.

This project was funded by CDC grants ELC/EIP HAI NU50CK000490-05-11, ELC AMD NU50CK000508-05-00, and ELC ARI NU50CK000508-05-00.

## REFERENCES

1. Centers for Disease Control and Prevention, 2024. *Clostridiodides difficile* infection (CDI) surveillance. US Department of Health and Human Services, Centers for Disease Control and Prevention. https://www.cdc.gov/healthcare-associated-infections/php/haic-eip/cdiff.html

2. Centers for Disease Control and Prevention, 2019. Antibiotic resistance threats in the United States, 2019. US Department of Health and Human Services, Centers for Disease Control and Prevention.

3. Buddle, J.E. and Fagan, R.P., 2023. Pathogenicity and virulence of *Clostridioides difficile*. Virulence, 14(1), p.2150452.

4. Song, J.H. and Kim, Y.S., 2019. Recurrent *Clostridium difficile* infection: risk factors, treatment, and prevention. Gut and liver, 13(1), p.16.

5. McDonald, L.C., Coignard, B., Dubberke, E., Song, X., Horan, T., Kutty, P.K. and Ad Hoc *Clostridium difficile* Surveillance Working Group, 2007. Recommendations for surveillance of *Clostridium difficile*–associated disease. Infection Control & Hospital Epidemiology, 28(2), pp.140–145.

6. Durovic, A., Widmer, A.F., Frei, R. and Tschudin-Sutter, S., 2017. Distinguishing Clostridium difficile recurrence from reinfection: independent validation of current recommendations. infection control & hospital epidemiology, 38(8), pp.891–896.

7. Barbar, R., Brazelton, J.N., Carroll, K.C., Lewis, S., Bourdas, D., Tembo, A., Gluck, L., Hakim, H. and Hayden, R.T., 2023. Molecular epidemiology and genetic relatedness of *Clostridioides difficile* isolates in pediatric oncology and transplant patients using whole genome sequencing. Clinical Infectious Diseases, 76(3), pp.e1071–e1078.

8. Kumar, N., Miyajima, F., He, M., Roberts, P., Swale, A., Ellison, L., Pickard, D., Smith, G., Molyneux, R., Dougan, G. and Parkhill, J., 2016. Genome-based infection tracking reveals dynamics of Clostridium difficile transmission and disease recurrence. Clinical Infectious Diseases, 62(6), pp.746–752.

9. Knight, D.R., Imwattana, K., Collins, D.A., Lim, S.C., Hong, S., Putsathit, P. and Riley, T.V., 2023. Genomic epidemiology and transmission dynamics of recurrent *Clostridioides difficile* infection in Western Australia. European Journal of Clinical Microbiology & Infectious Diseases, 42(5), pp.607–619.

10. Cho, J., Cunningham, S., Pu, M., Lennon, R.J., Dens Higano, J., Jeraldo, P., Sampathkumar, P., Shannon, S., Kashyap, P.C. and Patel, R., 2021. *Clostridioides difficile* whole-genome sequencing differentiates relapse with the same strain from reinfection with a new strain. Clinical Infectious Diseases, 72(5), pp.806–813.

11. Kulecka, M., Waker, E., Ambrozkiewicz, F., Paziewska, A., Skubisz, K., Cybula, P., Targoński, Ł., Mikula, M., Walewski, J. and Ostrowski, J., 2021. Higher genome variability within metabolism genes associates with recurrent *Clostridium difficile* infection. BMC microbiology, 21, pp.1–10.

12. Didelot, X., Eyre, D.W., Cule, M., Ip, C.L., Ansari, M.A., Griffiths, D., Vaughan, A., O’Connor, L., Golubchik, T., Batty, E.M. and Piazza, P., 2012. Microevolutionary analysis of *Clostridium difficile* genomes to investigate transmission. Genome biology, 13, pp.1–13.

13. He, M., Miyajima, F., Roberts, P., Ellison, L., Pickard, D.J., Martin, M.J., Connor, T.R., Harris, S.R., Fairley, D., Bamford, K.B. and D’Arc, S., 2013. Emergence and global spread of epidemic healthcare-associated *Clostridium difficile*. Nature genetics, 45(1), pp.109–113.

14. Eyre, D.W., Cule, M.L., Wilson, D.J., Griffiths, D., Vaughan, A., O’Connor, L., Ip, C.L., Golubchik, T., Batty, E.M., Finney, J.M. and Wyllie, D.H., 2013. Diverse sources of *C. difficile* infection identified on whole-genome sequencing. New England Journal of Medicine, 369(13), pp.1195–1205.

15. Norsigian, C.J., Danhof, H.A., Brand, C.K., Oezguen, N., Midani, F.S., Palsson, B.O., Savidge, T.C., Britton, R.A., Spinler, J.K. and Monk, J.M., 2020. Systems biology analysis of the *Clostridioides difficile* core-genome contextualizes microenvironmental evolutionary pressures leading to genotypic and phenotypic divergence. NPJ systems biology and applications, 6(1), p.31.

16. MALDI Biotyper CA System User Manual, Revision H, January 2019. Burker Daltonics, Inc.

17. Andrews, S., 2010. FastQC: a quality control tool for high throughput sequence data. Available online. https://www.bioinformatics.babraham.ac.uk/projects/fastqc/.

18. Wood, D.E., Lu, J. and Langmead, B., 2019. Improved metagenomic analysis with Kraken 2. Genome biology, 20, pp.1–13.

19. Seemann, T., 2018. Shovill: faster SPAdes (or better SKESA/Megahit/Velvet) assembly of Illumina reads. https://github.com/tseemann/shovill

20. Gurevich, A., Saveliev, V., Vyahhi, N. and Tesler, G., 2013. QUAST: quality assessment tool for genome assemblies. Bioinformatics, 29(8), pp.1072–1075.

21. Jolley, K.A. and Maiden, M.C., 2010. BIGSdb: scalable analysis of bacterial genome variation at the population level. BMC bioinformatics, 11, pp.1–11.

22. Seemann, T., 2018. MLST. https://github.com/tseemann/mlst

23. Seemann, T., 2014. Prokka: rapid prokaryotic genome annotation. Bioinformatics, 30(14), pp.2068–2069.

24. Page, A.J., Cummins, C.A., Hunt, M., Wong, V.K., Reuter, S., Holden, M.T., Fookes, M., Falush, D., Keane, J.A. and Parkhill, J., 2015. Roary: rapid large-scale prokaryote pan genome analysis. Bioinformatics, 31(22), pp.3691–3693.

25. Minh, B.Q., Schmidt, H.A., Chernomor, O., Schrempf, D., Woodhams, M.D., Von Haeseler, A. and Lanfear, R., 2020. IQ-TREE 2: new models and efficient methods for phylogenetic inference in the genomic era. Molecular biology and evolution, 37(5), pp.1530–1534.

26. Seemann T. 2015. Snippy: fast bacterial variant calling from NGS reads. https://github.com/tseemann/snippy.

27. Wick, R.R., Judd, L.M., Gorrie, C.L. and Holt, K.E., 2017. Unicycler: resolving bacterial genome assemblies from short and long sequencing reads. PLoS computational biology, 13(6), p.e1005595.

28. Kolmogorov, M., Yuan, J., Lin, Y. and Pevzner, P.A., 2019. Assembly of long, error-prone reads using repeat graphs. Nature biotechnology, 37(5), pp.540–546.

29. Seeman T., 2014. ABRicate. https://github.com/tseemann/abricate

30. Johansson, M.H., Bortolaia, V., Tansirichaiya, S., Aarestrup, F.M., Roberts, A.P. and Petersen, T.N., 2021. Detection of mobile genetic elements associated with antibiotic resistance in *Salmonella enterica* using a newly developed web tool: MobileElementFinder. Journal of Antimicrobial Chemotherapy, 76(1), pp.101–109.

31. Liu, M., Li, X., Xie, Y., Bi, D., Sun, J., Li, J., Tai, C., Deng, Z. and Ou, H.Y., 2019. ICEberg 2.0: an updated database of bacterial integrative and conjugative elements. Nucleic acids research, 47(D1), pp.D660–D665.

32. Robertson, J. and Nash, J.H., 2018. MOB-suite: software tools for clustering, reconstruction and typing of plasmids from draft assemblies. Microbial genomics, 4(8), p.e000206.

33. Team, R.C., 2020. R: A language and environment for statistical computing, R Foundation for Statistical Computing.

34. Cinelli, C., Ferwerda, J. and Hazlett, C., 2020. sensemakr: Sensitivity analysis tools for OLS in R and Stata. Available at SSRN 3588978.

35. Dayananda, P. and Wilcox, M.H., 2019. A review of mixed strain *Clostridium difficile* colonization and infection. Frontiers in microbiology, 10, p.692.

36. Hell, M., Permoser, M., Chmelizek, G., Kern, J.M., Maass, M., Huhulescu, S., Indra, A. and Allerberger, F., 2011. *Clostridium difficile* infection: monoclonal or polyclonal genesis?. Infection, 39(5), pp.461–465.

37. Miles-Jay, A., Snitkin, E.S., Lin, M.Y., Shimasaki, T., Schoeny, M., Fukuda, C., Dangana, T., Moore, N., Sansom, S.E., Yelin, R.D. and Bell, P., 2023. Longitudinal genomic surveillance of carriage and transmission of *Clostridioides difficile* in an intensive care unit. Nature Medicine, 29(10), pp.2526–2534.

38. Evans, D.R., Griffith, M.P., Sundermann, A.J., Shutt, K.A., Saul, M.I., Mustapha, M.M., Marsh, J.W., Cooper, V.S., Harrison, L.H. and Van Tyne, D., 2020. Systematic detection of horizontal gene transfer across genera among multidrug-resistant bacteria in a single hospital. Elife, 9, p.e53886.

39. Evans, D., Sundermann, A., Griffith, M., Srinivasa, V.R., Mustapha, M., Chen, J., Dubrawski, A., Cooper, V., Harrison, L. and Van Tyne, D., 2023. Empirically derived sequence similarity thresholds to study the genomic epidemiology of plasmids shared among healthcare-associated bacterial pathogens. EBioMedicine, 93.

40. Yaffe, E. and Relman, D.A., 2020. Tracking microbial evolution in the human gut using Hi-C reveals extensive horizontal gene transfer, persistence and adaptation. Nature microbiology, 5(2), pp.343–353.

41. Durrant, M.G., Li, M.M., Siranosian, B.A., Montgomery, S.B. and Bhatt, A.S., 2020. A bioinformatic analysis of integrative mobile genetic elements highlights their role in bacterial adaptation. Cell host & microbe, 27(1), pp.140–153.

42. Abante, J., Wang, P.L. and Salzman, J., 2023. DIVE: a reference-free statistical approach to diversity-generating and mobile genetic element discovery. Genome Biology, 24(1), p.240.

43. Acman, M., van Dorp, L., Santini, J.M. and Balloux, F., 2020. Large-scale network analysis captures biological features of bacterial plasmids. Nature communications, 11(1), p.2452.

44. Arredondo-Alonso, S., Willems, R.J., Van Schaik, W. and Schürch, A.C., 2017. On the (im) possibility of reconstructing plasmids from whole-genome short-read sequencing data. Microbial Genomics, 3(10), p.e000128.

